# Socioeconomic position, adverse childhood experiences, and menstrual symptoms in two generations of a prospective UK cohort

**DOI:** 10.64898/2026.08.27.26361513

**Authors:** Gemma Sawyer, Bushra Farooq, Kate Birnie, Abigail Fraser, Deborah A. Lawlor, Gemma C. Sharp, Laura D. Howe

## Abstract

**Background:** Inequalities exist for many health outcomes, but there is limited evidence regarding menstrual symptoms despite their importance for health and wellbeing. We aimed to investigate inequalities in menstrual symptoms according to socioeconomic position and childhood adversity.

**Methods:** In two generations (G0 mothers and G1 offspring) from the Avon Longitudinal Study of Parents and Children (ALSPAC), a UK prospective cohort study, we examined associations of multiple indicators of socioeconomic position (SEP) and adverse childhood experiences (ACEs) with menstrual symptoms (pain, abnormal uterine bleeding, and premenstrual syndrome (PMS) measured 3-8-years post-birth in G0 and 17-21-years-old in G1), using multivariable logistic regression. Samples ranged from 4,828 to 9,335 G0 participants and 1,288 to 2,757 G1 participants depending on the exposure-outcome association. Missing data were addressed using multiple imputation and inverse probability weighting.

**Results:** Financial difficulties were associated with greater odds of menstrual pain (G1 OR 1.41; 95% CI 1.07, 1.86: G0 OR 1.55; 95% CI 1.36, 1.76) and irregular cycles (G1 OR 1.60; 95% CI 1.12, 2.29: G0 OR 1.48; 95% CI 1.27, 1.72) in both generations, as well as with short/long cycle lengths in G0 only. Lower education and manual social class were also associated with these three menstrual symptoms in at least one generation. Conversely, higher SEP was associated with PMS in both generations. Higher cumulative ACEs were consistently associated with menstrual pain (4+ compared to none: G1 OR 2.15; 95% CI 1.48, 3.11: G0 OR 1.52; 95% CI 1.29, 1.80) and irregular cycles (G1 OR 1.92; 95% CI 1.20, 3.09: G0 OR 1.54; 95% CI 1.26, 1.87) but not cycle length. Lower parental education, financial difficulties, and cumulative ACEs were associated with heavy bleeding in G1 offspring only, whereas financial difficulties, own manual social class, and cumulative ACEs were associated with prolonged bleeding in G0 mothers only. Higher cumulative ACEs were also associated with PMS in G1 offspring only.

**Conclusions:** We found evidence of inequalities according to socioeconomic disadvantage and childhood adversity for multiple menstrual symptoms, although some associations were only observed in one generation. Findings suggest that menstrual symptoms are disproportionately experienced by socially and socioeconomically disadvantaged women.

## Background

Menstrual symptoms (including menstrual pain (dysmenorrhea), abnormal uterine bleeding (AUB), and premenstrual syndrome (PMS)) affect large proportions of girls, women, and people who menstruate, with impacts on health and wellbeing (1–5). Certain demographic groups may be differentially likely to experience menstrual symptoms. Previous cross-sectional studies have demonstrated a higher prevalence of menstrual pain, heavy bleeding, prolonged bleeding, abnormal cycle lengths, and PMS amongst those with lower incomes or recent financial problems (6–9). Irregular and prolonged bleeding, abnormal cycle length, and PMS have also been shown to be more prevalent among individuals with lower levels of education in cross-sectional studies (6,9–11). However, these findings have not been consistent, with other studies showing no relationship between income and pain or irregular bleeding (9–12), or between education and pain, cycle length, or PMS (6,9).

These inconsistencies may reflect the modest size of most studies as well as differences in setting, participant age, and assessment of menstrual symptoms. These studies are also weakened by their cross-sectional design, either minimal confounder adjustment or over-adjustment for variables on the causal pathway between SEP and menstrual symptoms (i.e., mediators) such as physical activity or depression, and the use of selected samples (such as university students) who tend to be more affluent than the general population. Addressing these weaknesses is crucial to better understand potential inequalities in menstrual symptoms.

Evidence of the relationship between adverse childhood experiences (ACEs) and menstrual symptoms is more consistent, with studies showing associations between abuse, neglect, and household dysfunction and the prevalence and severity of PMS (13–18). A systematic review of cross-sectional and case-control studies examining associations between childhood adversity and pelvic pain concluded that higher cumulative ACE exposure was associated with menstrual pain in adolescence and early adulthood (19). Although the authors reported inconsistent evidence for the relationship between specific individual ACEs (i.e., sexual abuse, parental separation, and parental death) and pain, they attributed this to methodological limitations, primarily over-adjusting for mediators.

Several studies have also demonstrated associations between overall ACE exposure and irregular and heavy bleeding (20,21). The associations of other types of abnormal uterine bleeding (AUB; such as prolonged bleeding and short/long cycle lengths) have been less studied. A systematic review of cross-sectional studies exploring stressful events in adulthood, including sexual harassment and incarceration, demonstrated a link with AUB, justifying further investigation into childhood exposures (22). Therefore, whilst evidence is indicative of associations between adversity and menstrual symptoms, the number of studies remains scarce, particularly for AUB, and the methodological quality is limited by cross-sectional designs and either no adjustment for confounders or over-adjustment for mediators such as smoking or alcohol use.

This study aims to examine how exposure to lower socioeconomic position (SEP; indexed with education, occupation, and financial difficulties) and ACEs (total and individual) are associated with six menstrual symptom outcomes in mothers and daughters from a UK population-based longitudinal cohort.

## Methods

### Participants

Pregnant women resident in Avon, UK with expected delivery dates between April 1991 and December 1992 were invited to take part in the Avon Longitudinal Study of Parents and Children (ALSPAC). 14,203 unique women (generation 0; G0) were initially enrolled in the study with 14,541 pregnancies. 13,988 children (generation 1; G1) were alive at age 1 year. ALSPAC has been described in detail elsewhere (23,24). Please note that the study website contains details of all the data that is available through a fully searchable data dictionary and variable search tool: http://www.bristol.ac.uk/alspac/researchers/our-data/. G0 participants with relevant exposure, covariate, and outcome data, as well as G1s assigned female at birth, are included here. As the outcomes were measured at different timepoints and/or required additional exclusions, there were four analysis samples for the G1 daughters (participant flowchart in Figure 1, referred to as G1 from here on) and three for the G0 mothers (Figure 2, referred to as G0 from here on).

**Figure 1.**
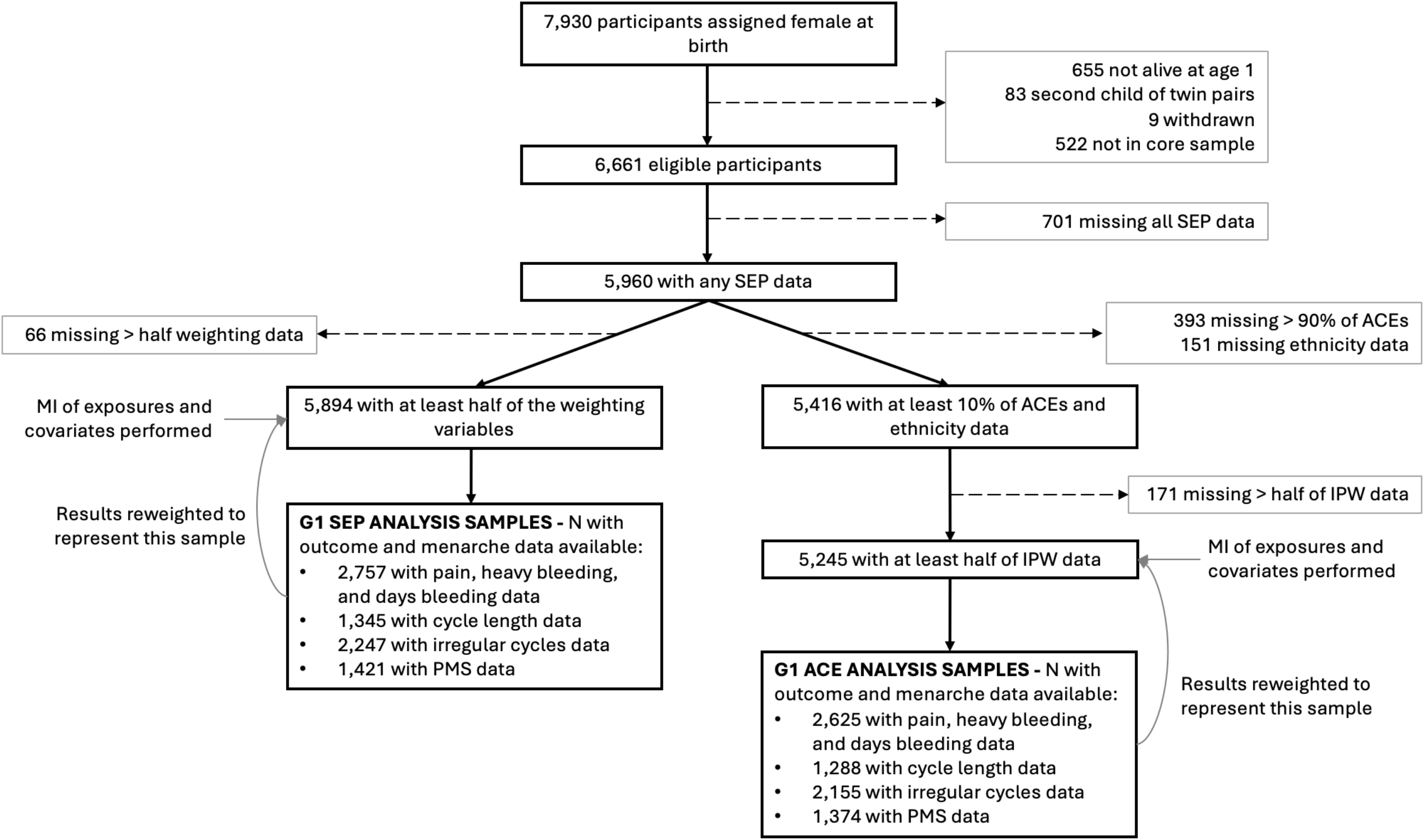
Flow diagram of the G1(Generation 1) offspring participants into the current study samples. Abbreviations: SEP, socioeconomic position; ACE, adverse childhood experience; MI, multiple imputation; PMS, premenstrual syndrome.

**Figure 2.**
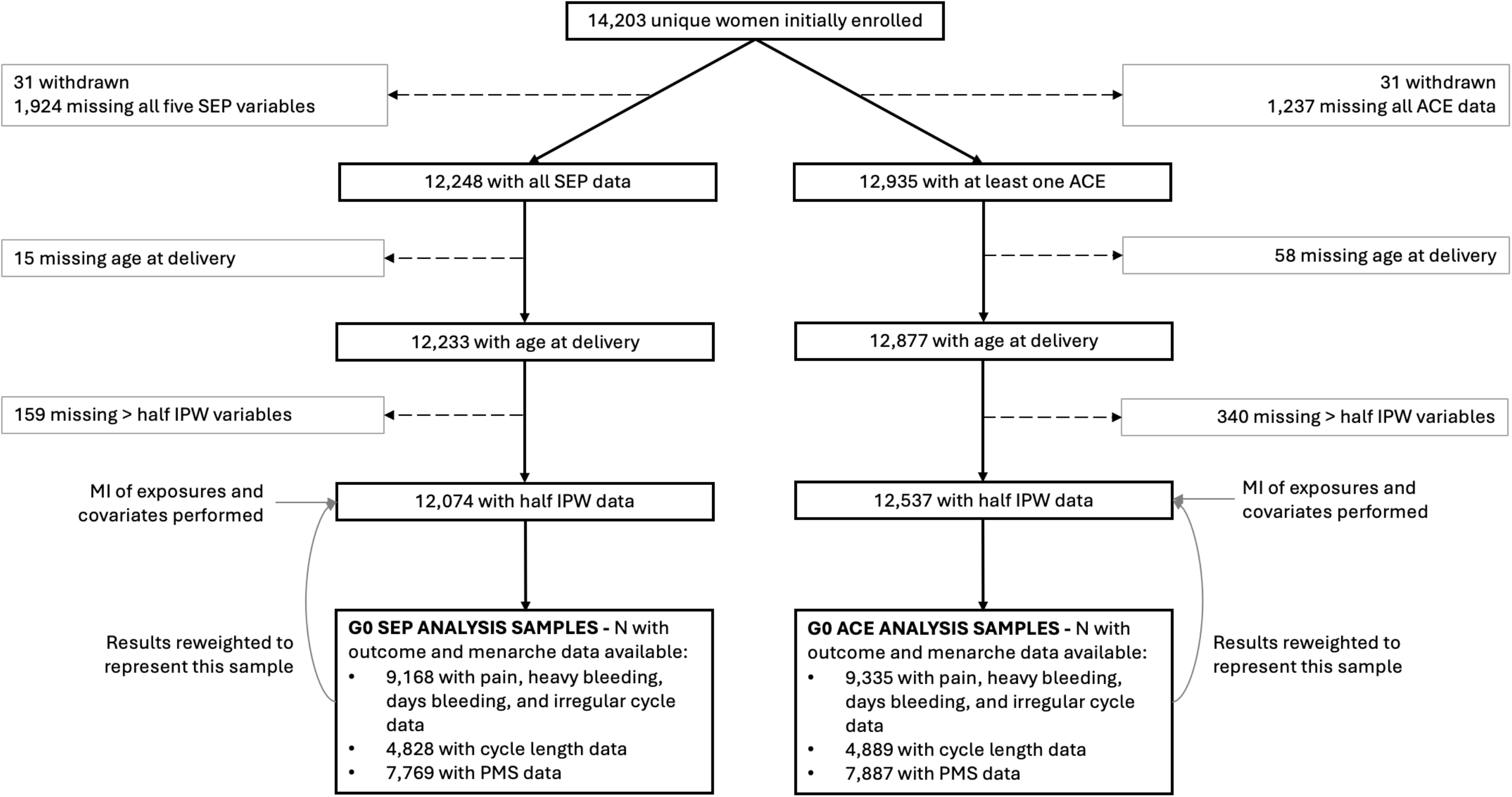
Flow diagram of the G0 (Generation 0) women participants into the current study samples. Abbreviations: SEP, socioeconomic position; ACE, adverse childhood experience; MI, multiple imputation; PMS, premenstrual syndrome.

### Exposures

#### G1 SEP measures

Parental measures reflecting knowledge-, resource-, and class-related aspects of SEP were self-reported by G0 via postal questionnaires at 32 weeks’ gestation. These included highest parental education (‘CSE (certificate of secondary education) or vocational (lowest)’, ‘O (ordinary) level’, ‘A (advanced) level’, and ‘degree (highest)’), highest parental occupational social class (‘manual’ or ‘non-manual’ determined by the 1991 British Office of Population and Census Statistics classification system), and financial difficulties during pregnancy (‘severe difficulties affording food, clothing, heating, accommodation, or things for the baby’ or ‘mild/no difficulties affording all items’).

### G0 SEP measures

Adulthood SEP in G0 was defined based upon their highest education, highest occupational social class, and financial difficulties during their pregnancy. In addition, G0 childhood SEP was indexed according to their parents’ highest education and highest occupational social class. All measures were self-reported by G0 at 32 weeks’ gestation and categorised in the same way as the G1 measures.

### G1 ACE measures

Derivation of ACEs in G1 has been described in detail previously (25). Ten ACEs experienced between birth and age 11 (preceding menarche for 93.3% of participants) were included for G1. These included physical, emotional, and sexual abuse, emotional neglect, violence between parents, parental mental health problems, household substance abuse, parental separation, bullying, and parental conviction. ACEs were derived from 302 individual items from mother-, partner-, and child-completed questionnaires, reported both prospectively and retrospectively reported (Supplementary Table 1) (26,27). Each ACE was binary (‘present’ or ‘absent’) and derived if participants had at least 50% non-missing items. A cumulative ACE variable (categorised as ‘none’, ‘one’, ‘two’, ‘three’, or ‘four or more’ ACEs) was also derived.

### G0 ACE measures

G0 ACEs have been derived previously (28). Eight ACEs (physical, emotional, and sexual abuse, emotional neglect, violence between parents, parental mental health problems, household substance abuse, parental separation; note that analysis of bullying and parental conviction was not possible due to no/limited data) were included for G0 between birth and age 18. 24 individual items contributing to these ACEs were all retrospectively self-reported via questionnaires during pregnancy (12 and 32 weeks’ gestation) and 33 months post-birth (Supplementary Table 2). As there were fewer individual measures contributing to each ACE here compared with G1, only one non-missing relevant item was required to derive each ACE. As with G1, we derived a cumulative ACE variable (categorised as ‘none’, ‘one’, ‘two’, ‘three’, or ‘four or more’ ACEs).

### Menstrual Symptom Outcomes

Both generations were asked about menstruation in a range of retrospective questionnaires and clinic assessments (described in detail in Supplementary Note 1). (29)

#### G1 daughters

At 16- or 17-years-old, menstrual-related pain (‘severe cramps’ or ‘no severe cramps’), heavy or prolonged bleeding (‘yes’ or ‘no’), prolonged bleeding (‘≥7 days’ or ‘<7 days’), and cycle length (‘abnormal: <24 or >38 days’ or ‘normal: 24-38 days’) were measured. Irregular bleeding was measured at 17.8 years clinic (‘irregular’ or ‘regular’) and PMS-related symptoms were measured at age 21 (‘any symptom: very fatigued, irritable, anxious, depressed, other’ or ‘no symptoms’).

### G0 mothers

Menstrual-related pain (‘very or moderate’ or ‘mild or not at all’), heavy bleeding (‘very or moderate’ or ‘mild or not at all’), prolonged bleeding (‘≥7 days’ or ‘<7 days’), and irregular bleeding (‘very or moderate’ or ‘mild or not at all’) were measured in one of four questionnaires (earliest non-missing response from 2.8, 3.9, 5.1, and 6.1 years post-birth).

PMS-related symptoms were measured at 6.1 years post-birth and cycle length was measured at 8.1 years post-birth.

### Confounders

Confounders were selected that could plausibly cause exposure and outcome (DAGs shown in Supplementary Figures 1 and 2). Ethnicity (‘white’ or ‘non-white’; G0-reported at 32 weeks’ gestation) was included as a confounder in all models and, when ACEs were the exposure, parental SEP variables were also included. Due to the relationship between age and parity, baseline age (years) was adjusted for in G0 to reduce the age-related variation in menstrual symptom outcomes. In G0, we also report results additionally adjusting for age at menarche (years; self-reported at 12 week’s gestation, 8.1, or 11.2 years post-birth) when own adulthood SEP and ACEs were the exposures. This adjustment was not necessary when parental SEP was the exposure nor for any G1 exposure as age at menarche would represent a mediator (i.e. on the causal pathway from exposure to outcome) here instead of a confounder.

## Statistical analysis

All analyses were conducted in Stata (version 18.0). Multivariable logistic regression models were used to estimate the association between each exposure and each menstrual symptom outcome, adjusting for relevant confounders. When exposures were categorical (i.e., three or more levels), a likelihood ratio (LR) test was conducted to compare a continuous with a categorical exposure model. P values from the continuous model are presented where no statistical difference was indicated, whereas p values from a post-estimation Wald test are presented when there was evidence for a statistical difference (Supplementary Table 3).

We also conducted a number of sensitivity analyses (details are provided in Supplementary Note 2). Briefly, these involved alternative exposures (parent-specific SEP due to different maternal and paternal effects being shown previously (30)), alternative outcomes (symptom severity and separating heavy from prolonged bleeding), and excluding certain groups (<three years since menarche due to possible changes in menstrual symptoms after the transition from anovulatory to ovulatory cycles and hormonal contraceptive users due to possible amelioration of symptoms following initiation). Analysis code is available on GitHub at https://github.com/GemmaS17.

### Missing data

We used multiple imputation (MI) to impute exposure and covariate data, but not outcome data due to uncertainty about whether available variables could adequately predict missing menstrual symptom values. To address possible selection bias as a result of loss to follow-up in completion of the menstrual symptom questionnaires, we conducted inverse probability weighting (IPW) alongside MI to reweight the analytic sample back to a sample closer to the fully eligible sample based on the probability of having non-missing outcome data (Supplementary Figure 3). Variables predictive of non-missing outcomes were based on previous studies (Supplementary Note 3 and Tables 4 and 5) and imputed using MI (31,32). A separate MI procedure was conducted for G1 and G0, as well as the SEP exposure analyses and the ACE exposure analyses. We required participants to have a minimum amount of exposure data (one SEP variable / 10% of ACEs data in G1 or one ACE variable in G0 due to differences in the number of ACE items contributing to each generation) and half of the weighting variables (as well as age at delivery in G0). All analytic variables were included in MI equations; however, as we did not wish to utilise imputed menstrual symptom outcomes in the analysis, these imputed values were deleted post-imputation. Other auxiliary variables were included for MI of the ACEs data to strengthen the missing at random assumption (Supplementary Tables 6 and 7). Details of the number of imputations and iterations are provided in Supplementary Note 4. Following MI, we calculated results using a within approach, which conducts both analysis steps related to IPW in each imputed dataset (calculating the probability of non-missing outcome data and estimating the exposure-outcome association with the calculated probabilities) before pooling using Rubin’s rules. Statistical packages *mi impute chained* in Stata and *mice* in R were used.

The prevalence/means of each variable before and after MI are presented in Supplementary Tables 8 and 9. Sensitivity analyses were conducted including truncated weights at the 95^th^ and 99^th^ percentiles (to assess if results were driven by few participants with large weights) and an alternative MI procedure with no outcome imputation (to assess whether bias was induced through outcome imputation and deletion) (33,34).

## Results

### Descriptives

Amongst G1 (Ns ranged from 1,288 to 2,757 depending on the exposure-outcome association), menstrual-related pain was the most commonly reported menstrual symptom, whereas PMS-related symptoms and heavy bleeding were the most common symptom reported by G0 (Ns ranged from 4,828-9,335) (Tables 1 and 2; see Supplementary Tables 10 and 11 for the proportions of exposures and confounders according to each menstrual symptom).

**Table 1.**
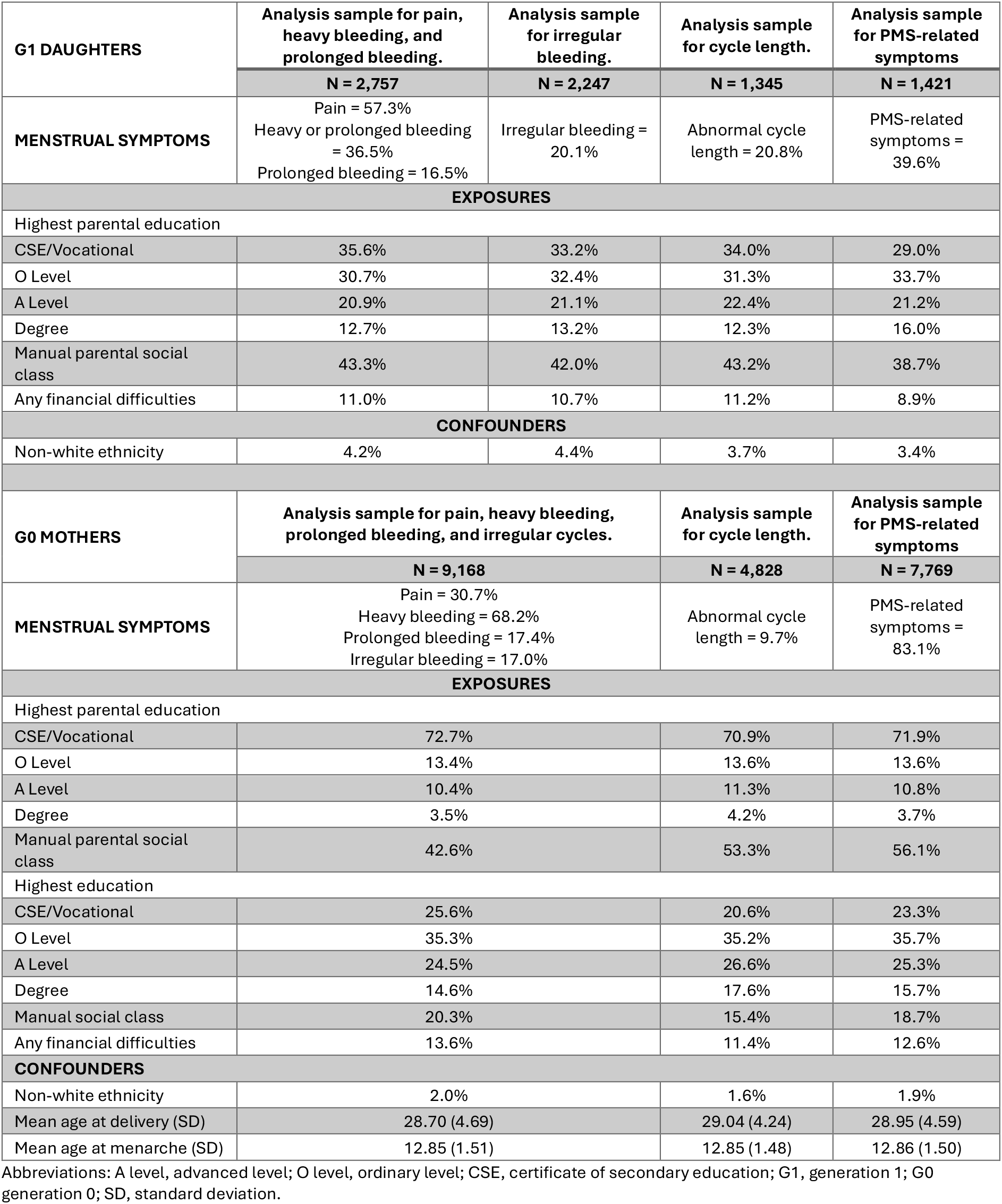
Descriptive statistics for exposure, outcomes, and confounders in the different analytical samples exploring the associations between socioeconomic position and menstrual symptoms in G1 daughters and G0 mothers.

**Table 2.**
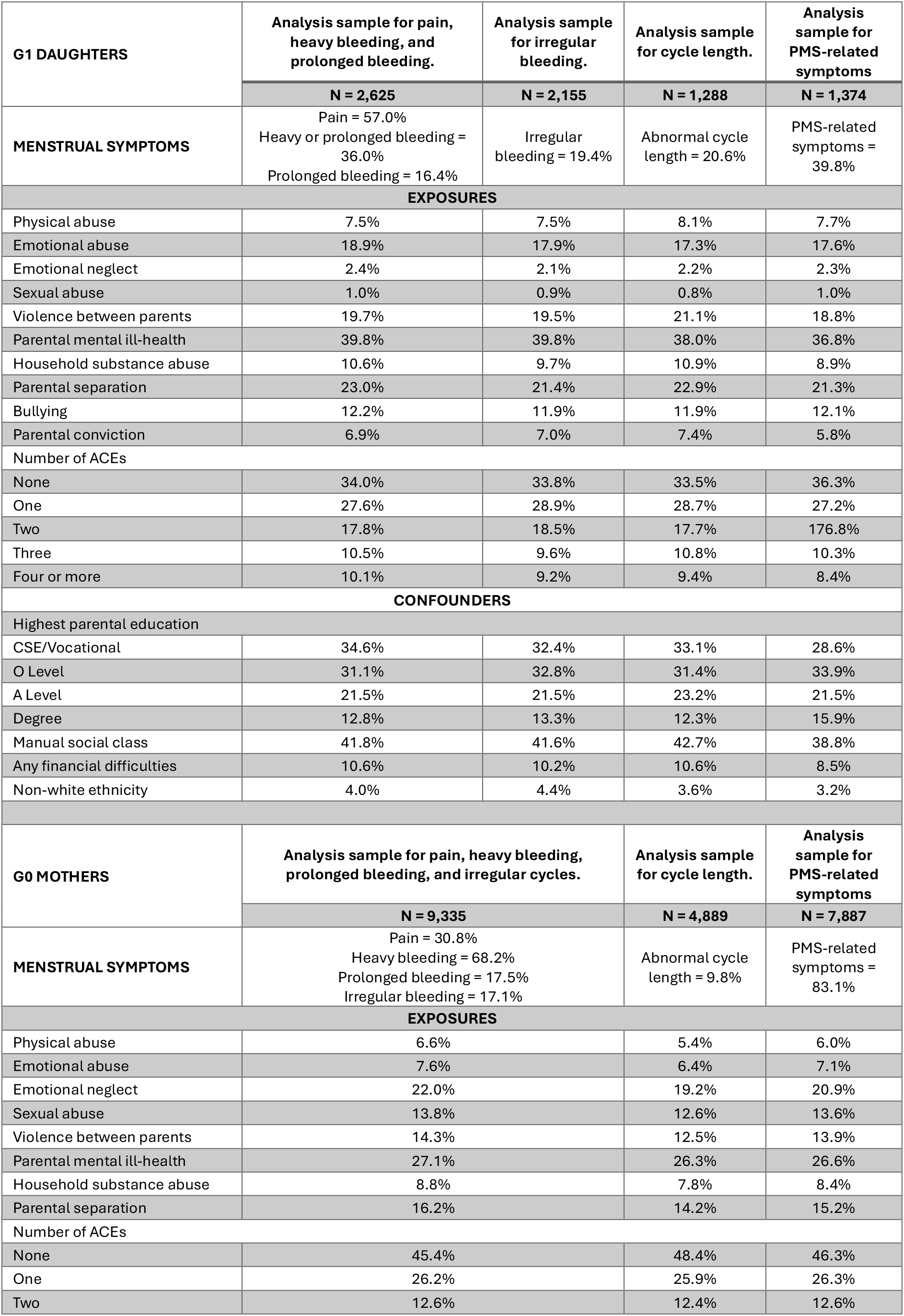
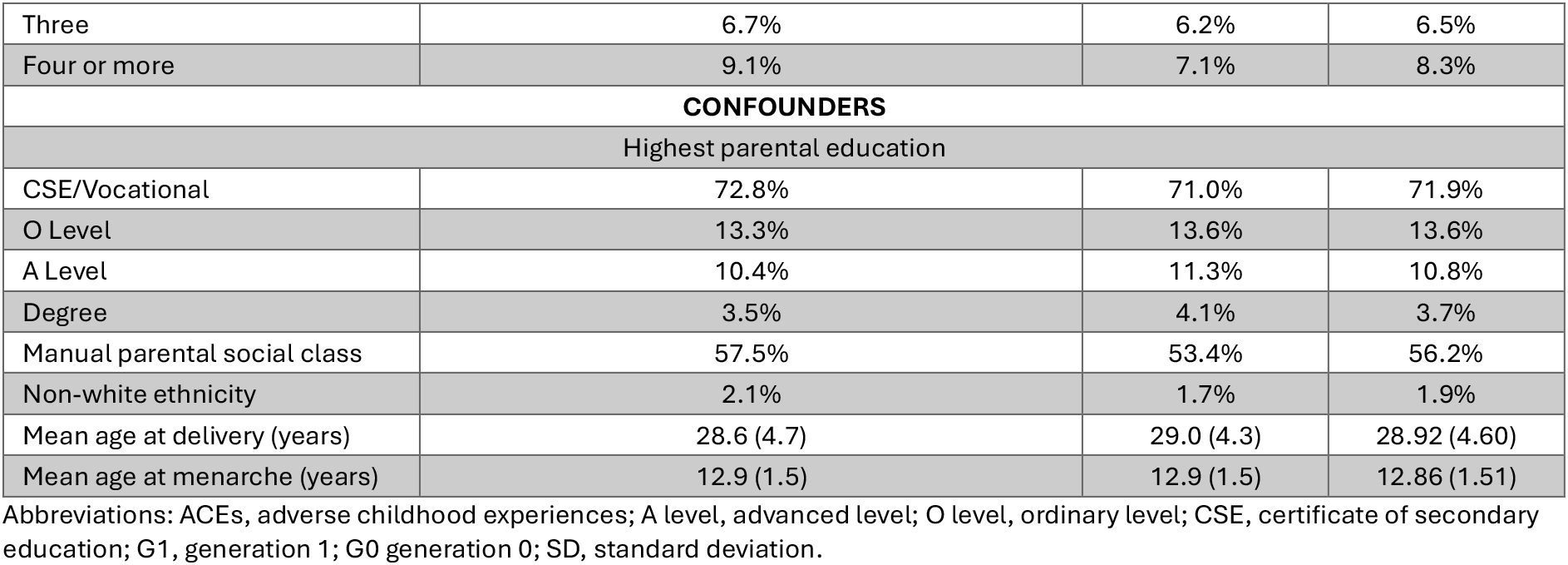
Descriptive statistics for exposure, outcomes, and confounders in the different analytical samples exploring the associations between adverse childhood experiences and menstrual symptoms in G1 daughters and G0 mothers.

| G1 DAUGHTERS | Analysis sample for pain, heavy bleeding, and prolonged bleeding. | Analysis sample for irregular bleeding. | Analysis sample for cycle length. | Analysis sample for PMS-related symptoms |
| --- | --- | --- | --- | --- |
|  | N = 2,625 | N = 2,155 | N = 1,288 | N = 1,374 |
| MENSTRUAL SYMPTOMS | Pain = 57.0%<br>Heavy or prolonged bleeding = 36.0%<br>Prolonged bleeding = 16.4% | Irregular bleeding = 19.4% | Abnormal cycle length = 20.6% | PMS-related symptoms = 39.8% |
| EXPOSURES |  |  |  |  |
| Physical abuse | 7.5% | 7.5% | 8.1% | 7.7% |
| Emotional abuse | 18.9% | 17.9% | 17.3% | 17.6% |
| Emotional neglect | 2.4% | 2.1% | 2.2% | 2.3% |
| Sexual abuse | 1.0% | 0.9% | 0.8% | 1.0% |
| Violence between parents | 19.7% | 19.5% | 21.1% | 18.8% |
| Parental mental ill-health | 39.8% | 39.8% | 38.0% | 36.8% |
| Household substance abuse | 10.6% | 9.7% | 10.9% | 8.9% |
| Parental separation | 23.0% | 21.4% | 22.9% | 21.3% |
| Bullying | 12.2% | 11.9% | 11.9% | 12.1% |
| Parental conviction | 6.9% | 7.0% | 7.4% | 5.8% |
| Number of ACEs |  |  |  |  |
| None | 34.0% | 33.8% | 33.5% | 36.3% |
| One | 27.6% | 28.9% | 28.7% | 27.2% |
| Two | 17.8% | 18.5% | 17.7% | 17.8% |
| Three | 10.5% | 9.6% | 10.8% | 10.3% |
| Four or more | 10.1% | 9.2% | 9.4% | 8.4% |
| CONFOUNDERS |  |  |  |  |
| Highest parental education |  |  |  |  |
| CSE/Vocational | 34.6% | 32.4% | 33.1% | 28.6% |
| O Level | 31.1% | 32.8% | 31.4% | 33.9% |
| A Level | 21.5% | 21.5% | 23.2% | 21.5% |
| Degree | 12.8% | 13.3% | 12.3% | 15.9% |
| Manual social class | 41.8% | 41.6% | 42.7% | 38.8% |
| Any financial difficulties | 10.6% | 10.2% | 10.6% | 8.5% |
| Non-white ethnicity | 4.0% | 4.4% | 3.6% | 3.2% |
| G0 MOTHERS | Analysis sample for pain, heavy bleeding, prolonged bleeding, and irregular cycles. |  | Analysis sample for cycle length. | Analysis sample for PMS-related symptoms |
|  | N = 9,335 |  | N = 4,889 | N = 7,887 |
| MENSTRUAL SYMPTOMS | Pain = 30.8%<br>Heavy bleeding = 68.2%<br>Prolonged bleeding = 17.5%<br>Irregular bleeding = 17.1% |  | Abnormal cycle length = 9.8% | PMS-related symptoms = 83.1% |
| EXPOSURES |  |  |  |  |
| Physical abuse | 6.6% |  | 5.4% | 6.0% |
| Emotional abuse | 7.6% |  | 6.4% | 7.1% |
| Emotional neglect | 22.0% |  | 19.2% | 20.9% |
| Sexual abuse | 13.8% |  | 12.6% | 13.6% |
| Violence between parents | 14.3% |  | 12.5% | 13.9% |
| Parental mental ill-health | 27.1% |  | 26.3% | 26.6% |
| Household substance abuse | 8.8% |  | 7.8% | 8.4% |
| Parental separation | 16.2% |  | 14.2% | 15.2% |
| Number of ACEs |  |  |  |  |
| None | 45.4% |  | 48.4% | 46.3% |
| One | 26.2% |  | 25.9% | 26.3% |
| Two | 12.6% |  | 12.4% | 12.6% |

|  |  |  |  |
| --- | --- | --- | --- |
| Three | 6.7% | 6.2% | 6.5% |
| Four or more | 9.1% | 7.1% | 8.3% |
| **CONFOUNDERS** |  |  |  |
| Highest parental education |  |  |  |
| CSE/Vocational | 72.8% | 71.0% | 71.9% |
| O Level | 13.3% | 13.6% | 13.6% |
| A Level | 10.4% | 11.3% | 10.8% |
| Degree | 3.5% | 4.1% | 3.7% |
| Manual parental social class | 57.5% | 53.4% | 56.2% |
| Non-white ethnicity | 2.1% | 1.7% | 1.9% |
| Mean age at delivery (years) | 28.6 (4.7) | 29.0 (4.3) | 28.92 (4.60) |
| Mean age at menarche (years) | 12.9 (1.5) | 12.9 (1.5) | 12.86 (1.51) |

Abbreviations: ACEs, adverse childhood experiences; A level, advanced level; O level, ordinary level; CSE, certificate of secondary education; G1, generation 1; G0 generation 0; SD, standard deviation.

### Associations between SEP and menstrual symptoms

The overall pattern of results indicates that lower SEP (parental for G1 and own for G0) was associated with a greater likelihood of reporting menstrual pain, irregular bleeding, and abnormal cycle lengths in both generations, as well as a lower likelihood of PMS-related symptoms. Differences were seen between generations regarding heavy and prolonged bleeding as low SEP was associated with heavy bleeding in daughters only but with prolonged bleeding in mothers only. The results, adjusting for confounders (ethnicity in G1; ethnicity and age at delivery and age at menarche when own SEP is the exposure in G0), are outlined in more detail below and presented in Figure 3. Additional adjustment for age at menarche in G0 did not alter results. Full results are presented in Supplementary Table 12 and 13.

**Figure 3.**
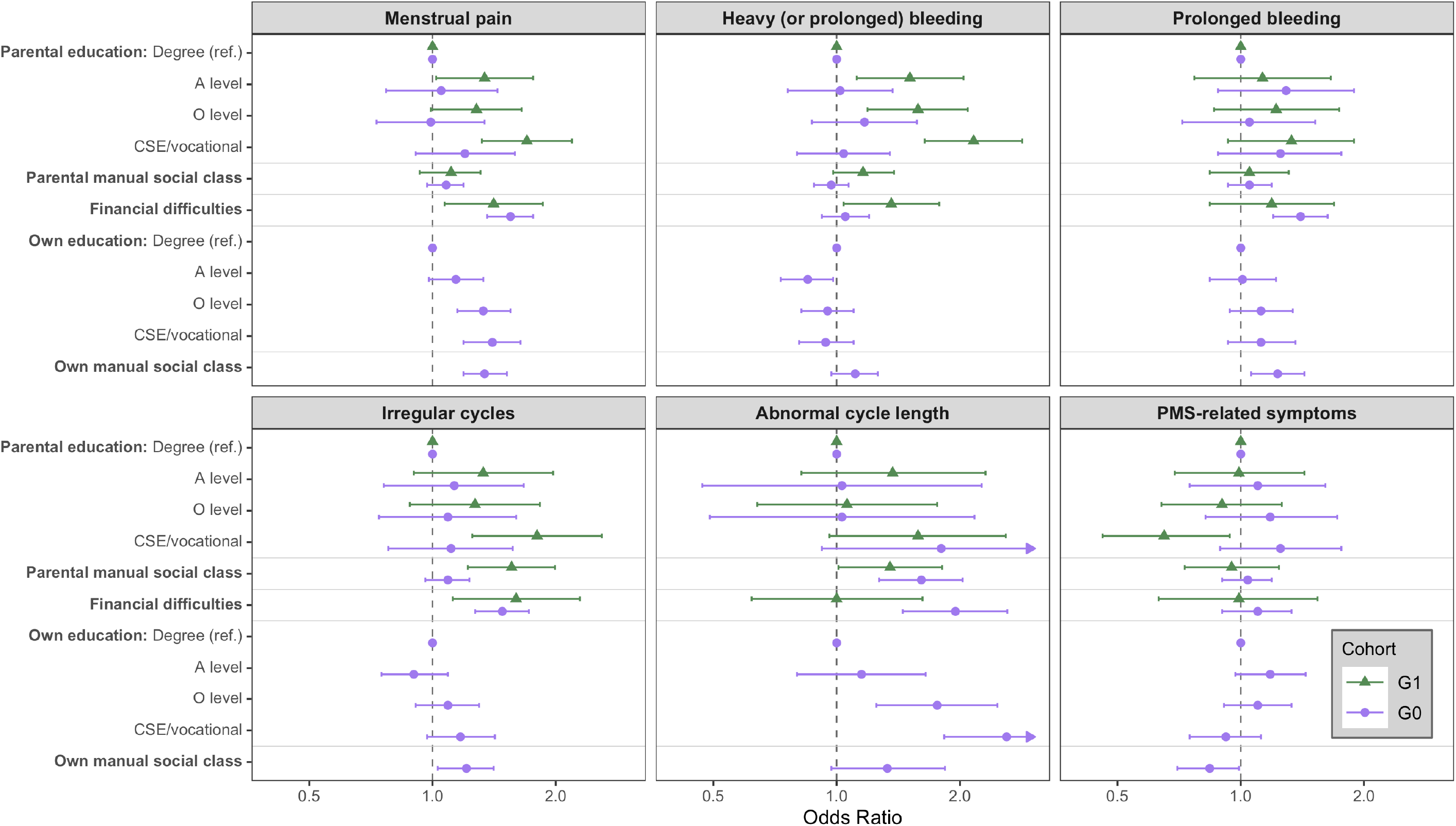
Multivariable logistic regression analysis of the associations between socioeconomic position and menstrual symptoms in two generations. G1 estimates adjusted for ethnicity: participant Ns were 2,757 for pain, heavy or prolonged bleeding, and prolonged bleeding; 2,247 for irregular cycles; 1,345 for cycle length; and 1,421 for PMS-related symptoms. G0 estimates adjusted for ethnicity and age at delivery, plus age at menarche for own exposures: participant Ns were 9,168 for pain, heavy bleeding, prolonged bleeding, and irregular cycles; 4,828 for cycle length; and 7,769 for PMS-related symptoms. Abbreviations: A level, advanced level; O level, ordinary level; CSE, certificate of secondary education; G1, generation 1; G0 generation 0.

### Menstrual pain

Parental financial difficulties (OR 1.41; 95% CI 1.07, 1.86) during pregnancy and lower parental education, in a dose-response manner (compared to degree: A level 1.34; 95% CI 1.02, 1.76: O level 1.28; 95% CI 0.99, 1.65: CSE/vocational 1.70; 95% CI 1.32, 2.19), were associated with higher odds of menstrual pain in G1. In G0, financial difficulties (OR 1.55; 95% CI 1.36, 1.76), lower own education (e.g., CSE/vocational v degree OR 1.40; 95% CI 1.19, 1.64), and manual occupations (OR 1.34; 95% CI 1.19, 1.52) were linked to greater odds of menstrual pain.

### Heavy (or prolonged) bleeding

In G1, parental financial difficulties before birth (OR 1.36; 95% CI 1.04, 1.78) and lower parental education (compared to degree: A level 1.51, 95% CI 1.12, 2.04: O level 1.58, 95% CI 1.19, 2.09: CSE/vocational 2.16; 95% CI 1.64, 2.84) were associated with higher odds of heavy bleeding. In G0, SEP measures showed little consistent association with heavy bleeding.

### Prolonged bleeding

There was little evidence that any SEP measure was associated with prolonged bleeding in G1. In G0, however, own manual occupations (OR 1.23; 95% CI 1.06, 1.43) and financial difficulties (OR 1.40; 95% CI 1.20, 1.63) were associated with a greater likelihood of prolonged bleeding.

### Cycle length

Parental manual social class was associated with abnormal cycle lengths in both G1 (OR 1.35; 95% CI, 1.01, 1.81) and G0 (OR 1.61; 95% CI 1.27, 2.04). Additionally, financial difficulties (OR 1.95; 95% CI 1.45, 2.61), lower parental education (CSE/vocational v degree OR 1.81; 95% CI 0.93, 3.53), and lower own education (CSE/vocational v degree OR 2.60; 95% CI 1.83, 3.70) were associated with greater odds of abnormal cycle lengths in G0.

### Irregular cycles

All SEP measures examined in G1 were associated with a greater likelihood of irregular cycles, including lower parental education (CSE/vocational v degree OR 1.80; 95% CI 1.25, 2.59), parental manual social class (OR 1.56; 95% CI 1.22, 1.99), and financial difficulties (OR 1.60; 95% CI 1.12, 2.29). In G0, own manual occupation (OR 1.21; 95% CI 1.03, 1.41) and financial difficulties during pregnancy (OR 1.48; 95% CI 1.27, 1.72) were associated with higher odds of irregular bleeding.

### PMS-related symptoms

Parental education was the only SEP measure associated with PMS-related symptoms in the G1 cohort, with lower odds as education decreased (compared to degree: A level 0.99; 95% CI 0.69, 1.43: O level 0.90; 95% CI 0.64, 1.26: CSE/vocational level 0.65; 95% 0.46, 0.94). Own manual social class, but no other SEP measure, was associated with a lower likelihood of PMS in G0 (OR 0.84, 95% CI 0.70, 0.99).

### Associations between ACEs and menstrual symptoms

Results indicate that greater ACE exposure (both in terms of cumulative and many individual ACEs) was associated with menstrual pain and irregular bleeding in both generations. Similar to the SEP results, experiencing more ACEs was associated with heavy bleeding in the daughters only and with prolonged bleeding in the mothers only. There was little consistent evidence that ACEs were associated with PMS-related symptoms or abnormal cycle lengths. Adjusted results (adjusting for ethnicity and parental SEP, plus age at delivery and age at menarche in G0) are outlined below in more detail and presented in Figure 4 (cumulative ACEs) and Table 3 (individual ACEs). Full results are available in Supplementary Table 14 and 15.

**Figure 4.**
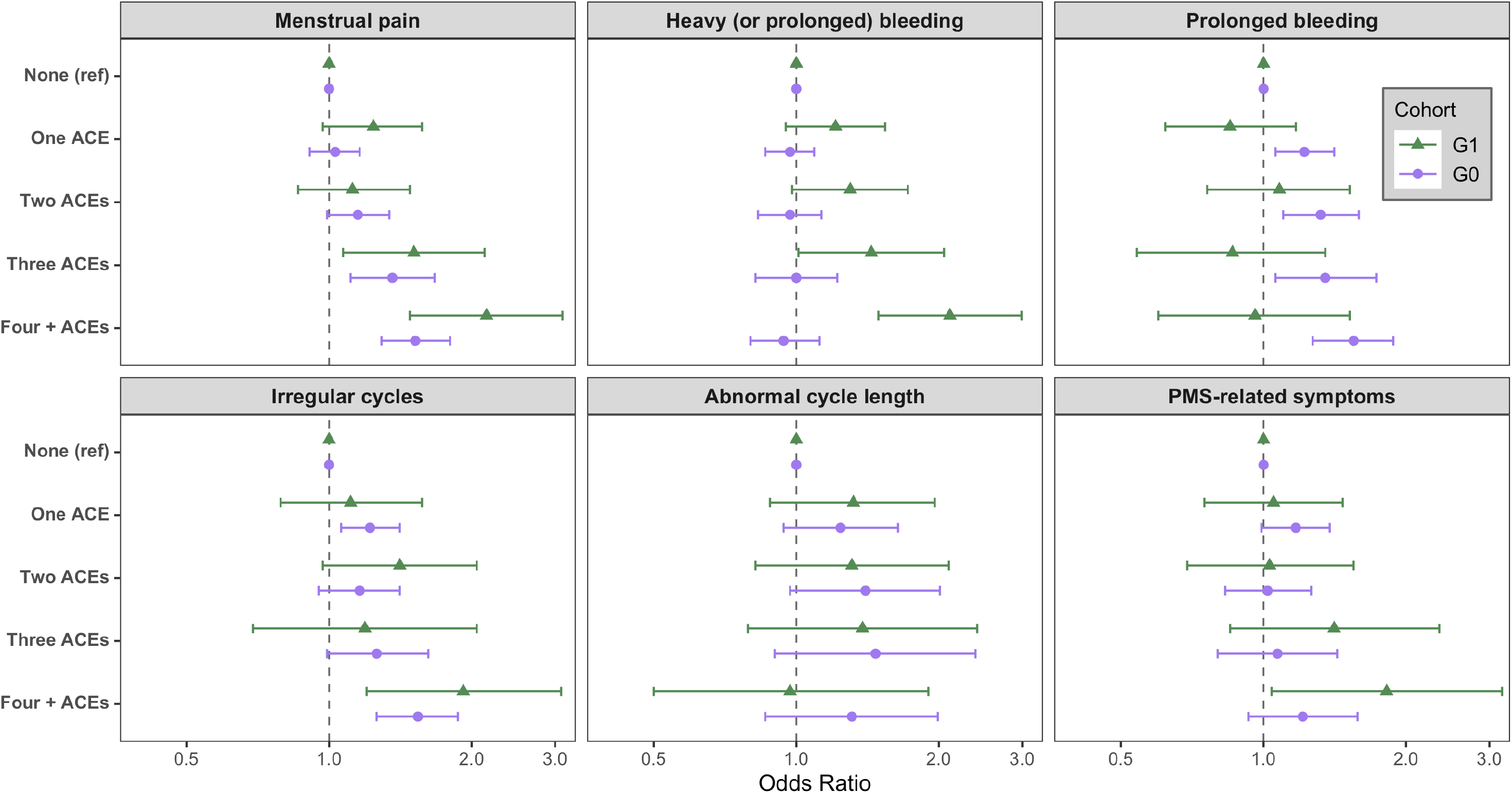
Multivariable logistic regression analysis of the associations between cumulative adverse childhood experiences and menstrual symptoms in two generations. G1 estimates adjusted for ethnicity, parental education, parental social class, and parental financial difficulties: participant Ns were 2,625 for pain, heavy or prolonged bleeding, and prolonged bleeding; 2,155 for irregular cycles; 1,288 for cycle length; and 1,374 for PMS-related symptoms. G0 estimates adjusted for ethnicity, age at delivery, age at menarche, parental education, and parental social class: participant Ns were 9,335 for pain, heavy bleeding, prolonged bleeding, and irregular cycles; 4,889 for cycle length; and 7,887 for PMS-related symptoms. Abbreviations: ACE; adverse childhood experience; G1, generation 1; G0 generation 0.

**Table 3.** Multivariable logistic regression analysis of the associations between individual adverse childhood experiences and menstrual symptoms in two generations.

| EXPOSURE | COHORT | Menstrual pain | Heavy (or prolonged bleeding) | Prolonged bleeding | Irregular cycles | Abnormal cycle length | PMS-related symptoms |
| --- | --- | --- | --- | --- | --- | --- | --- |
| Physical abuse | G1 Offspring | 1.51 (1.03, 2.21)<br>p = 0.035 | 1.43 (0.96, 2.11)<br>p = 0.077 | 1.04 (0.64, 1.71)<br>p = 0.866 | 1.22 (0.73, 2.02)<br>p = 0.449 | 0.91 (0.49, 1.68)<br>p = 0.762 | 1.39 (0.80, 2.42)<br>p = 0.239 |
|  | G0 Women | 1.41 (1.19, 1.68)<br>p = <0.001 | 1.02 (0.85, 1.22)<br>p = 0.845 | 1.43 (1.17, 1.75)<br>p = 0.001 | 1.32 (1.07, 1.63)<br>p = 0.009 | 1.15 (0.73, 1.81)<br>p = 0.550 | 1.00 (0.76, 1.30)<br>p = 0.974 |
| Emotional abuse | G1 Offspring | 1.54 (1.19, 1.98)<br>p = 0.001 | 1.28 (1.00, 1.63)<br>p = 0.046 | 0.90 (0.63, 1.27)<br>p = 0.538 | 0.88 (0.59, 1.30)<br>p = 0.507 | 1.54 (1.02, 2.34)<br>p = 0.041 | 1.23 (0.83, 1.82)<br>p = 0.297 |
|  | G0 Women | 1.49 (1.27, 1.76)<br>p = <0.001 | 1.02 (0.86, 1.21)<br>p = 0.830 | 1.33 (1.09, 1.61)<br>p = 0.005 | 1.27 (1.04, 1.55)<br>p = 0.020 | 1.01 (0.65, 1.56)<br>p = 0.974 | 1.36 (1.04, 1.78)<br>p = 0.023 |
| Emotional neglect | G1 Offspring | 0.84 (0.46, 1.54)<br>p = 0.568 | 1.78 (0.98, 3.24)<br>p = 0.060 | 1.26 (0.62, 2.57)<br>p = 0.520 | 0.70 (0.27, 1.83)<br>p = 0.471 | 1.43 (0.54, 3.78)<br>p = 0.469 | 1.17 (0.44, 3.12)<br>p = 0.752 |
|  | G0 Women | 1.38 (1.23, 1.54)<br>p = <0.001 | 1.06 (0.95, 1.18)<br>p = 0.320 | 1.31 (1.15, 1.49)<br>p = <0.001 | 1.27 (1.11, 1.45)<br>p = 0.001 | 1.02 (0.75, 1.37)<br>p = 0.915 | 1.16 (0.98, 1.37)<br>p = 0.088 |
| Sexual abuse | G1 Offspring | 2.57 (0.77, 8.54)<br>p = 0.124 | 1.38 (0.48, 4.02)<br>p = 0.549 | 0.69 (0.16, 2.97)<br>p = 0.614 | 0.75 (0.17, 3.44)<br>p = 0.715 | 0.77 (0.14, 4.31)<br>p = 0.765 | 1.65 (0.39, 7.02)<br>p = 0.496 |
|  | G0 Women | 1.23 (1.08, 1.40)<br>p = 0.002 | 1.01 (0.88, 1.16)<br>p = 0.866 | 1.27 (1.08, 1.48)<br>p = 0.004 | 1.34 (1.14, 1.57)<br>p = <0.001 | 1.06 (0.76, 1.48)<br>p = 0.732 | 0.98 (0.81, 1.19)<br>p = 0.866 |
| Violence between parents | G1 Offspring | 1.29 (1.01, 1.66)<br>p = 0.043 | 1.22 (0.94, 1.58)<br>p = 0.136 | 0.89 (0.63, 1.25)<br>p = 0.494 | 1.32 (0.90, 1.94)<br>p = 0.149 | 0.53 (0.31, 0.90)<br>p = 0.019 | 1.22 (0.84, 1.76)<br>p = 0.299 |
|  | G0 Women | 1.32 (1.14, 1.52)<br>p = <0.001 | 1.03 (0.90, 1.19)<br>p = 0.654 | 1.33 (1.13, 1.57)<br>p = 0.001 | 1.21 (1.02, 1.43)<br>p = 0.025 | 1.58 (1.15, 2.19)<br>p = 0.005 | 0.95 (0.77, 1.16)<br>p = 0.602 |
| Parental mental health problems | G1 Offspring | 1.50 (1.25, 1.81)<br>p = <0.001 | 1.50 (1.24, 1.82)<br>p = <0.001 | 1.14 (0.89, 1.46)<br>p = 0.286 | 1.11 (0.84, 1.45)<br>p = 0.463 | 1.06 (0.77, 1.45)<br>p = 0.729 | 1.23 (0.93, 1.64)<br>p = 0.153 |
|  | G0 Women | 1.13 (1.02, 1.25)<br>p = 0.017 | 0.96 (0.87, 1.06)<br>p = 0.452 | 1.23 (1.09, 1.39)<br>p = 0.001 | 1.16 (1.03, 1.32)<br>p = 0.016 | 0.98 (0.77, 1.25)<br>p = 0.881 | 1.27 (1.09, 1.48)<br>p = 0.002 |
| Household substance abuse | G1 Offspring | 1.29 (0.93, 1.78)<br>p = 0.129 | 1.17 (0.85, 1.62)<br>p = 0.333 | 0.59 (0.36, 0.99)<br>p = 0.046 | 1.62 (1.04, 2.53)<br>p = 0.033 | 0.79 (0.45, 1.39)<br>p = 0.419 | 1.37 (0.83, 2.28)<br>p = 0.220 |
|  | G0 Women | 1.21 (1.03, 1.43)<br>p = 0.020 | 0.88 (0.75, 1.03)<br>p = 0.122 | 1.17 (0.96, 1.42)<br>p = 0.119 | 1.11 (0.91, 1.36)<br>p = 0.303 | 0.90 (0.57, 1.40)<br>p = 0.637 | 0.88 (0.69, 1.11)<br>p = 0.276 |
| Parental separation | G1 Offspring | 1.03 (0.82, 1.29)<br>p = 0.788 | 1.18 (0.94, 1.48)<br>p = 0.165 | 0.87 (0.64, 1.19)<br>p = 0.386 | 1.57 (1.15, 2.13)<br>p = 0.005 | 1.07 (0.73, 1.56)<br>p = 0.740 | 0.97 (0.68, 1.39)<br>p = 0.865 |
|  | G0 Women | 1.06 (0.93, 1.20)<br>p = 0.378 | 0.85 (0.75, 0.96)<br>p = 0.007 | 1.05 (0.91, 1.23)<br>p = 0.493 | 1.16 (1.00, 1.35)<br>p = 0.049 | 1.70 (1.29, 2.24)<br>p = <0.001 | 0.97 (0.81, 1.16)<br>p = 0.719 |
| Bullying* | G1 Offspring | 1.18 (0.88, 1.58)<br>p = 0.272 | 1.46 (1.11, 1.93)<br>p = 0.007 | 1.57 (1.10, 2.24)<br>p = 0.013 | 1.53 (1.06, 2.21)<br>p = 0.024 | 1.43 (0.89, 2.28)<br>p = 0.137 | 1.38 (0.90, 2.11)<br>p = 0.134 |
| Parental conviction* | G1 Offspring | 1.42 (0.94, 2.15)<br>p = 0.092 | 1.43 (0.94, 2.17)<br>p = 0.093 | 0.96 (0.56, 1.65)<br>p = 0.883 | 1.65 (0.97, 2.80)<br>p = 0.065 | 0.78 (0.39, 1.56)<br>p = 0.486 | 1.67 (0.86, 3.26)<br>p = 0.133 |

#### Menstrual pain

Higher number of ACEs were associated with greater odds of menstrual pain in both G1 (compared to no ACE: one 1.24; 95% CI 0.97, 1.57: two 1.12; 95% CI 0.86, 1.48: three 1.51; 95% CI 1.07, 2.13: four or more 2.15; 95% CI 1.48, 3.11) and G0 (one 1.03; 95% CI 0.91, 1.16: two 1.15; 95% CI 0.99, 1.34: three 1.36; 95% CI 1.11, 1.67: four or more 1.52; 95% CI 1.29, 1.80), with a clear dose-response pattern. Most individual ACEs were also associated with a higher likelihood of pain in both G1 and G0, with abuse-related ACEs showing the largest effect estimates.

### Heavy (or prolonged) bleeding

In G1, but not G0, higher cumulative ACE exposure was associated with higher odds of reporting heavy or prolonged bleeding (four or more v none OR 2.11; 95% CI 1.49, 2.99). Bullying and parental mental health problems were associated with greater odds of heavy or prolonged bleeding in G1 and the effect estimates for emotional abuse, physical abuse, and emotional neglect were indicative of the same association; however, CIs were wide. There was little evidence that individual ACEs were associated with heavy bleeding in G0.

### Prolonged bleeding

In G1, only two individual ACEs (bullying and household substance abuse) were associated with greater odds of prolonged bleeding. G0 results showed a different pattern, with greater cumulative ACE exposure (four or more v none OR 1.55; 95% CI 1.27, 1.88) and most individual ACEs being associated with higher odds of prolonged bleeding.

### Cycle length

Cumulative ACEs showed little association with the likelihood of abnormal cycle lengths in either G1 or G0 participants. Only one individual ACE in G1 (emotional abuse) and two individual ACEs in G0 (violence between parents and parental separation) were associated with higher odds of abnormal cycle length. G1 results were inconsistent, with violence between parents being associated with lower odds of abnormal lengths.

### Irregular cycles

Higher cumulative ACEs were associated with a greater likelihood of irregular cycles in both G1 (four or more v none OR 1.92; 95% CI 1.20, 3.09) and G0 (four or more v none OR 1.54; 95% CI 1.26, 1.87). Six of the eight individual ACEs examined in G0 were associated with a greater likelihood of reporting irregular bleeding, except for household substance abuse and parental separation. Notably, these two ACEs, plus bullying were associated with cycle irregularity in G1.

### PMS-related symptoms

In G1 (four or more v none OR 1.82; 95% CI 1.04, 3.19), but not G0, higher ACEs were associated with higher odds of PMS-related symptoms. No individual ACEs were associated with PMS in G1, whereas emotional abuse and parental mental health problems were associated with a greater likelihood of PMS-related symptoms in G0.

## Sensitivity analyses

Results from sensitivity analyses did not alter conclusions drawn from the main results (Supplementary Note 5 and Supplementary Tables 16-19). The results from analyses using truncated IPWs and the alternative MI procedure (i.e., no outcome imputation) were almost identical to the main results (Supplementary Tables 20 and 21).

## Discussion

This study found that low SEP and high ACE exposure was associated with higher odds of various menstrual symptoms in two generations of a population-based prospective cohort study. This pattern was most consistent for menstrual pain, irregular cycles, and abnormal cycle lengths (low SEP only), whereas results for heavy and prolonged bleeding were inconsistent between generations. The pattern for PMS-related symptoms was complex, tending to show evidence for associations between higher SEP and greater odds of PMS, but higher cumulative (but no individual) ACEs were also associated with PMS.

Comparison with previous cross-sectional literature is challenging due to the mixed results and limited number of studies, particularly for AUB. Evidence for the relationship between SEP and pain, irregular bleeding, and cycle length has been inconsistent, and discrepancies are likely due to methodological differences (i.e., symptom measurement, sample characteristics, and confounder adjustment) (6,7,9,10,12,30). For example, most studies have focused either on young people or broad reproductive-age samples without examining age differences, whereas our study specifically compares young individuals and those in their thirties at the time of symptom reporting. The few studies examining AUB have tended to support associations between low SEP and higher odds of heavy bleeding (consistent with the G1 findings) and prolonged bleeding (consistent with G0 findings) (6,7). Our finding that high SEP was associated with PMS broadly contradicts the few previous SEP studies that have reported either null associations or evidence for associations between lower SEP and PMS (6,11).

Our findings of higher odds of pain in people exposed to ACEs is consistent with previous systematic review evidence and a small number of studies have also supported the same relationship for irregular bleeding (consistent with G1 and G0 findings) and heavy bleeding (consistent with G1 findings only) (19–21). It is possible that the discrepancy with heavy bleeding for G0 (for both SEP and ACEs) is due to the higher prevalence of heavy bleeding (68% compared to 37% in the offspring). This may reflect different experiences of heavy bleeding, with many of the G0 case group experiencing heavy bleeding as a common feature of perimenopause, which may still have problematic consequences for people but reflects a different aetiology. However, divergences between offspring and mothers may also be due to differences in the measurement of ACEs as G1 ACEs covered those in early- and mid-childhood (birth to 11 years) and were predominantly prospectively reported by their mother whereas G0 ACEs were retrospectively self-reported across childhood (birth to 18 years). Different prevalences in the ACE exposures (e.g., emotional abuse is reported in 17-19% of G1 and 6-8% of G0) may therefore contribute to some of the differences reported between these generations.

Stressful events in adulthood have been linked with other AUB types and therefore the null findings regarding ACEs and cycle length, and ACEs and prolonged bleeding in G1, may be considered contradictory and it is unclear why these findings differ (22). Our current findings also contrast with a wide body of consistent evidence for associations between cumulative and individual ACEs across various domains with PMS-related symptoms (13–18). As with SEP, different PMS definitions may explain the inconsistency.

## Strengths and Limitations

A key strength of this study is its use of a prospective birth cohort spanning two generations (G0 mothers and G1 daughters), allowing assessment of how socioeconomic position and ACEs are associated with menstrual outcomes across generations. The use of a population-based prospective cohort also enabled inclusion of confounders without over-adjusting for mediators, a less selected sample than previous studies (e.g., university students or nurses), and reduced likelihood of reverse causality. However, despite use of MI and IPW to mitigate the impact of socially patterned attrition, ALSPAC remains more affluent and less diverse than the UK population. Underrepresented groups, such as non-white women, experiences of structural inequalities and discrimination likely intersect with their healthcare and menstrual experiences, possibly leading to different relationships than those currently reported. Also, although reverse causality is unlikely amongst G1 as exposures preceded menarche, this is not necessarily true for SEP and ACEs for G0. These exposures could potentially have been impacted by menstrual symptoms, although this is unlikely to fully account for the observed ACE associations.

Whilst it is also beneficial that we examined multiple aspects of SEP, the measures reflect a snapshot in time and, for G0, changes in working patterns around pregnancy may lead to fluctuations in occupation or income and younger participants may still be establishing their education or career. Examining both individual ACEs and an ACE score enables exploration of how different types of ACEs may be differentially associated with menstrual symptoms but also recognises that many ACEs co-occur, although this can incorrectly assume equivalence in those with the same score. Alternative methods, such as latent class analysis, can be used to define ACEs based on which experiences cluster together over time; however, these trajectories rarely replicate (35). Additionally, there may be some misclassification in G1 ACEs due to the items being predominantly prospectively reported by mothers, alongside recall bias in the retrospective self-reporting of G0 ACEs, as individuals may underreport or choose not to disclose adverse experiences on questionnaires, or may not recognise certain experiences as adverse until later in life. The use of multiple regression models increases the potential for a type I error; however, corrections for multiple testing would be over-stringent because the models are not independent due to inter-related and co-occurring aspects of SEP and childhood adversity and similarly co-occurring menstrual problems (36,37).

There are limitations regarding the measurement of menstrual symptoms, including their retrospective reporting (which may not agree with prospective reporting (38)) and their binary classification (removing variations in severity, chronicity, and underlying causes). We attempted to examine symptom severity (indexed through medical care seeking in G1 or self-reported severity in G0), with results tending to demonstrate either stronger associations in those with more severe symptoms or similar associations regardless of severity. However, this only partially addresses the heterogeneity, and future research should replicate analyses with detailed symptoms and compare idiopathic symptoms to those due to underlying conditions.

Similarly, clinical definitions could not be consistently utilised due to limitations with question wording, including the combination of heavy and prolonged bleeding in G1 and 7 days or more (instead of 9 or more) being the highest category for prolonged bleeding (3). Despite this, ALSPAC is one of the only large multigenerational studies with detailed prospective data on SEP, ACEs, and menstrual symptoms and thus is a unique and valuable cohort in which to examine the research question. New and ongoing cohort studies should endeavour to collect detailed, repeat data on menstrual experiences to allow replication.

## Possible mechanisms

There are several pathways that may explain the associations between SEP, ACEs, and menstrual symptoms. For example, structural and environmental factors that are patterned by SEP (such as access to nutritious food, safe spaces for physical activity, and exposure to tobacco and alcohol marketing) may shape health behaviours that have been linked with menstrual pain and AUB (1,10,30,39,40). Early life adversity and trauma have been linked with earlier pubertal timing, which, according to the accelerated life history theory, may be an adaptive biological response to environmental conditions; earlier age at menarche has in turn been linked with a range of menstrual symptoms (2,9,41–44). Adversity may also contribute to increased central nervous system sensitisation, leading to heightened pain perception (1,45–47). Psychosocial stress may operate via dysregulation of the hypothalamic-pituitary-adrenal (HPA) axis, which regulates complex hormone systems, and has been linked to inflammation, mood, and menstrual disorders (48–51). In addition, SEP and ACEs are associated with lower mental health and psychological wellbeing, which may interact with and possibly exacerbate menstrual symptoms (15,18,52,53).

## Future research and implications

In addition to future research exploring SEP and ACE trajectories and examining more detailed symptoms, it would also be beneficial to replicate current analyses in other, diverse, population-based cohorts. Currently, few cohorts have collected the necessary data on menstrual health to achieve this and, therefore, other sources of data may be worth exploring, including GP records and menstrual tracking apps. These have the potential to be representative, although social patterns in help-seeking behaviour and use of smartphones may limit this (54–57). Triangulation across different data sources would increase confidence in our conclusions and, as evidence builds, research should focus on identifying pathways involved in the associations to identify modifiable targets for intervention and reduce the burden of menstrual symptoms across the population, while addressing the disproportionate impacts that might be experienced by socially disadvantaged groups.

The highlighted inequalities in menstrual symptoms are particularly concerning given the wider adverse impacts such symptoms can have on broader health and wellbeing. For example, our previous research has demonstrated that pain and heavy bleeding are associated with more school absences and worse educational attainment, after adjusting for a range of confounders (58). This raises the possibility that menstrual symptoms may play a role in persistent socioeconomic disadvantage, both being more likely to be present in disadvantaged groups and more likely to restrict an individual’s ability to enhance their own education, and thus social position, highlighting the importance of improving menstrual health through a proportionate universalism approach to minimise such inequalities. More comprehensive menstrual education is required (in schools, medical training, and population-wide public health campaigns) to normalise conversations and reduce stigma, improve understanding and skills for management, empower individuals to identify symptoms and seek help, and reduce the likelihood of dismissal by healthcare services. The relationship between ACEs and menstrual symptoms may also justify the expansion of trauma informed care (TIC) practices in medical settings and schools; however, more evaluation is needed to ensure TIC has positive impacts and is not re-traumatising (59–62).

## Conclusion

This study demonstrates that lower SEP and greater number of ACEs are associated with menstrual-related pain, irregular bleeding, and abnormal cycle length (SEP only) in two generations from a population-based prospective cohort. Additionally, lower SEP and greater ACEs were associated with heavy bleeding in the daughters’ cohort and prolonged bleeding in the mothers’ cohort, which may be indicative of different relationships across the reproductive lifespan or across successive generations. Results were complex regarding PMS-related symptoms, possibly reflecting limitations in their definition and measurement. These findings identify specific subgroups at higher risk of menstrual symptoms, highlighting both the disproportionate burden of menstrual symptoms and the areas where research and interventions should be prioritised to reduce inequalities.

## List of abbreviations

A-level: Advanced Level
ACEs: Adverse Childhood Experiences
ALSPAC: Avon Longitudinal Study of Parents and Children
AUB: Abnormal Uterine Bleeding
CI: Confidence Interval
CSE: Certificate of Secondary Education
G0: Generation 0
G1: Generation 1
HPA: Hypothalamic-Pituitary Adrenal
IPW: Inverse Probability Weighting
MI: Multiple Imputation
O-level: Ordinary Level
OR: Odds Ratio
PMS: Premenstrual Syndrome
SEP: Socioeconomic Position
TIC: Trauma Informed Care
UK: United Kingdom

## Declarations

### Ethics approval and consent to participate

Ethical approval for the study was obtained from the ALSPAC Ethics and Law Committee and the Local Research Ethics Committees. Informed consent for the use of all data collected was obtained from participants following the recommendations of the ALSPAC Ethics and Law Committee at the time. Participants can contact the study team at any time to retrospectively withdraw consent for their data to be used. Study participation is voluntary and during all data collection sweeps, information was provided on the intended use of data. The completion of a questionnaire, either on paper or online, was considered to be written consent from participants to use their data for research purposes.

### Consent for publication

Not applicable

### Availability of data and materials

The informed consent obtained from ALSPAC (Avon Longitudinal Study of Parents and Children) participants does not allow the data to be made available through any third party maintained public repository. Supporting data are available from ALSPAC on request under the approved proposal number, B4175. Full instructions for applying for data access can be found here: http://www.bristol.ac.uk/alspac/researchers/access/. The ALSPAC study website contains details of all available data (http://www.bristol.ac.uk/alspac/researchers/our-data/). All analysis code utilised in this paper is available on GitHub at https://github.com/GemmaS17.

### Authors’ contributions

GS: Conceptualization, Methodology, Formal analysis, Writing – Original Draft, Visualization. GCS, LDH, AF, DAL: Conceptualization, Writing – Review & Editing, Supervision. BF, KB: Methodology, Writing – Review & Editing.

## Acknowledgements

We are extremely grateful to all the families who took part in this study, the midwives for their help in recruiting them, and the whole ALSPAC team, which includes data collection staff, data and administrations staff, technical managers and the technical staff with the Bristol Bioresource Laboratory, based within the University of Bristol. The informed consent obtained from ALSPAC (Avon Longitudinal Study of Parents and Children) participants does not allow the data to be made available through any third party maintained public repository. Supporting data are available from ALSPAC on request under the approved proposal number, B4175. Full instructions for applying for data access can be found here: http://www.bristol.ac.uk/alspac/researchers/access/. The ALSPAC study website contains details of all available data (http://www.bristol.ac.uk/alspac/researchers/our-data/).

