## Supplementary Material for "Socioeconomic position, adverse childhood experiences, and menstrual symptoms in two generations of a prospective UK cohort"

**ALSPAC SEP/ACEs Paper Supplement**

**Supplementary Note 1: Details of the menstrual symptom outcome measures**

The menstrual symptom outcomes in the G1 offspring were derived from responses to the following questions.

1. “Have you had any of the following symptoms associated with your period: severe cramps?”. Responses to this question at 16- or 17-years-old were used to derive a binary variable representing ‘menstrual-related pain’ or ‘not’.
2. At 16- or 17-years-old, participants were asked “Have you had any of the following symptoms associated with your period: heavy or prolonged bleeding?”. Participants could answer ‘yes’ or ‘no’ and these were used to derive a binary heavy or prolonged bleeding variable.
3. Participants were also asked “In the past year, how many days of bleeding have you usually had during each period?” at 16- or 17-years-old and were able to report the exact number of days bleeding or, if they were unsure, could select from one of three options: ‘3 days or less’, ‘4-6 days’, or ‘7 days or more’. To utilise all responses, a binary prolonged bleeding variable was derived reflecting ‘bleeding for 7 days or more’ or ‘bleeding for less than 7 days’.
4. “Are your periods regular?”, which was asked to participants at the 17.8-year clinic assessment, was used to derive a binary cycle regularity variables reflecting ‘irregular’ or ‘regular’ periods.
5. “If your periods are regular, how long on average would you say your cycle is? (i.e., number of days between each period e.g., 30, 28)”. Responses to this question at 16- or 17-years old were used to derive a binary cycle length variable reflecting ‘normative length between 24 and 38 days’ or ‘abnormal lengths below 24 or above 38 days’. Responses below 10 were excluded due to concerns that respondents had misunderstood the question and were reporting the number of days bleeding.
6. At age 21, participants were asked “Do you generally find that in the days before or during your periods you have particular problems” and “If yes, which problems do you experience: very fatigued, irritable, depressed, anxious, or other?”. Participants were able to indicate whether they experienced any of these symptoms (they were able to select more than one) before or during their period (or both). Responses were used to indicate whether participants reported ‘any symptom before or during their period’ or ‘no symptoms’.

Similar questions were presented to G0 women and used to derive the menstrual symptom outcomes in this study.

1. At four timepoints (2.8, 3.9, 5.1, and 6.1 years post-birth), participants were asked “How painful are your periods” and could response by selecting ‘very’, ‘moderately’, ‘mildly’, or ‘not at all’. A binary menstrual-related pain variable was derived representing ‘menstrual-related pain (very or moderately)’ or ‘no menstrual-related pain (mildly or not at all). The earliest response to any of the menstrual symptoms asked at these timepoints (see below) was utilised (i.e., if participants only responded to one question, the rest were set to missing).
2. “How heavy are your periods” was asked to participants at 2.8, 3.9, 5.1, and 6.1 years post-birth with response options including ‘very’, ‘moderately’, ‘mildly’, or ‘not at all’. Those who reported very or moderately were coded as experiencing ‘heavy bleeding’, whereas those who reported mildly or not at all were coded as ‘not experiencing heavy bleeding’.
3. Participants were also asked “How many days does bleeding usually last?” at the same four timepoints and were able to report the exact number of days. For consistency with G1 offspring, a binary prolonged bleeding variable was derived reflecting ‘bleeding for 7 days or more’ or ‘bleeding for less than 7 days’.
4. At one of four timepoints (2.8, 3.9, 5.1, and 6.1 years post-birth), participants were either asked “How would you describe your periods: irregular?” or “Are your periods irregular?” and were able to respond with ‘very’, ‘moderately’, ‘mildly’, or ‘not at all’. Responses were used to derive a binary cycle regularity variable representing ‘irregular (very or moderately)’ or ‘regular (mildly or not at all)’ periods.
5. “If regular, how many days were there from the start of one period to the start of the next one?”, which was presented to participants at 8.1-years post-birth (the earliest timepoint post-birth), was used to derive a binary cycle length variable reflecting ‘normative length between 24 and 38 days’ or ‘abnormal lengths below 24 or above 38 days’. Responses below 10 were excluded.
6. At 6.1-years post-birth (the earliest post-pregnancy timepoint), participants were asked “Do you generally find in the days before or during your periods you have particular problems: very fatigued, irritable, depressed, anxious, or other?”. As with G1 offspring, participants could select any and all symptoms, before or during their period, or both and responses were used to indicate whether participants reported ‘any symptom before or during their period’ or ‘no symptoms’.

**Supplementary Note 2: Sensitivity analyses**

A number of sensitivity analyses were conducted in both cohorts using observed data only.

1. To examine whether both parent’s SEP were associated with menstrual symptom in a similar manner, we estimated the associations between parent-specific (maternal and paternal/mother’s partner separately) SEP exposures and menstrual symptoms in both generations.
2. Due to the role hormonal contraception can play in impacting menstrual symptoms (often prescribed to ameliorate such symptoms), we excluded participants reporting hormonal contraception use (oral contraceptive pill only in G1 and oral contraceptive pill, intrauterine device, or coil in G0) when estimating the associations between exposures and outcomes.
3. To explore symptom severity, we included information on whether G1 offspring sought medical care for menstrual-related pain and heavy or prolonged bleeding (‘no symptom’, ‘symptom, but did not seek medical care’, or ‘symptom and sought medical care’). In G0 women, we utilised the original four-level response categories regarding pain, heavy bleeding, and irregular bleeding (‘very’, ‘moderately’, ‘mildly’, ‘not at all’).

We also conducted two additional sensitivity analyses in the G1 offspring only.

4. As there is some research indicating that menstrual symptoms in early reproductive life (up to an average of 3 years post-menarche) may be more severe due to not yet having transitioned from anovulatory to ovulatory menstruation, we excluded participants who reported on their menstrual symptoms within the first three years since menarche (this did not lead to any exclusions regarding PMS).
5. To separate heavy from prolonged bleeding, I estimated the associations between SEP and a four-level outcome variable which combined the heavy or prolonged variable and the prolonged bleeding variable (‘no heavy or prolonged bleeding and bleeding for <7 days’, ‘not heavy or prolonged bleeding but bleeding for ≥7 days’, ‘heavy or prolonged bleeding but bleeding for <7 days’ or ‘heavy or prolonged bleeding and bleeding for ≥7 days’).

**Supplementary Note 3: Variables for Inverse Probability Weighting (IPW)**

The variables that were considered to be used to develop IPWs predictive of non-missing outcome data were selected based on two previous ALSPAC studies (Cornish 2021, Cornish 2015) and are described in detail in Supplementary Table 4. Briefly, these included:

- Maternal age at delivery (continuous, 15 to 44 years);
- Maternal education (‘O level or lower’, ‘A level’, or ‘degree’, mother-reported during pregnancy);
- Maternal marital status (‘married’ or ‘not married’, mother-reported during pregnancy);
- Maternal smoking during pregnancy (‘any’ or ‘none’, mother-reported during pregnancy);
- Maternal ever smoking status (‘yes’ or ‘no’, mother-reported during pregnancy);
- Maternal depressive symptoms (continuous, 0 to 28, mother-reported during pregnancy based on the Edinburgh Postnatal Depression Scale (EPDS));
- Maternal parity (‘0’, ‘1’, ‘2+’, mother-reported during pregnancy);
- Maternal phone ownership (‘no working phone or incoming calls only’ or ‘working phone’, mother-reported during pregnancy);
- Maternal car ownership (‘yes’ or ‘no’, mother-reported during pregnancy);
- Crowding index (people in the house divided by number of rooms, ‘≤0.5’, ‘>0.5-0.75’, ‘>0.75-1’, or ‘>1’, mother-reported during pregnancy);
- Double glazing (‘any’ or ‘none’, mother-reported during pregnancy);
- Maternal age at first pregnancy (‘<20’, ’20-24’, ‘25+’, mother-reported during pregnancy);
- Financial difficulty score (continuous, 0 to 15, mother-reported during pregnancy);
- Family social class (‘manual’ or ‘non-manual’, mother-reported during pregnancy);
- Breastfeeding duration (‘never or <1 month’, ‘1 to <3 months’, ‘3 to <6 months’, or ‘6 months +’, mother-reported during early childhood); and
- Family adversity index (FAI; continuous, 0 to 18 reflecting the number of familial risk factors based on a range of variables from pregnancy and early childhood, when more than half are non-missing (Crawley 2012; Bowen 2005).

To select variables contributing to the IPW calculation, I conducted separate lasso predictive models for each outcome sample. In both cohorts, these identified similar predictive variables and so, to evaluate whether the same model could be used across all cohort samples, I compared an inclusive model (with all variables identified by the lasso model for at least one outcome) with the model identified by the lasso model for each outcome, using a LR test. The LR tests indicated no statistical difference between the models and, therefore, the same IPW model was used for each outcome (see Supplementary Table 5 information on the models considered).

**Supplementary Note 4: Multiple Imputation (MI) details**

The same approach was taken across almost all MIs to select the number of iterations and imputations. First, to establish the number of required iterations, I generated trace plots of the imputed means and SDs for each variable across 100 iterations. Examination of the trace plots allows identification of the required number of iterations for values to become stable. For the number of imputations, I initially determined this based on the percentage of missing data (i.e., 50 imputations when half of the sample are missing at least one variable) (White & Royston 2011). However, I conducted post-imputation checks to examine the performance of the MI and adjusted where necessary. Alongside simple comparisons between the proportions or means and distributions of observed with imputed data, I also assessed the fraction of missing information (FMI) and Monte Carlo error (MCE) estimates. The FMI is a parameter that reflects the amount of the total sampling variance that can be attributed to variation between the imputations, with a larger FMI (closer to 1 or 100%) indicating more variance as a result of missing data (UCLA Multiple imputation in stata; Madley-Dowd 2019). In general, the FMI decreases as estimates become more stable (usually with more imputations), and it is recommended that the FMI (as a percentage) be smaller than the number of imputations (UCLA again). Overall, this recommendation was satisfied, and I did not make any changes to the imputation number as a result. MCE tests are a method for assessing the statistical reproducibility of MI, with guidelines suggesting that (1) the MCE of coefficients should be <10% of the standard error, (2) the MCE of the test statistic should be <0.1, and (3) the MCE of p values should be <0.01 when the p value is 0.05 or <0.02 when the p value is 0.1 (White & Royston again). These criteria were satisfied for the majority of MI; however, some were violated regarding the initial MI procedure for the SEP analysis in G0 women, which resulted in me increasing the number of imputations. The MI procedure for the ACE analysis in G1 offspring adopted a slightly different approach as imputation of the exposure variables (ACEs) had been conducted previously. As such, I adopted the same approach as this previous work in terms of the auxiliary variables and number of iterations/imputations instead of originally determining these based on trace plots and proportions of missing data. I followed the same approach to check the MI performance and concluded no adjustments were required.

The final MI equations generated 50 imputed datasets across 50 iterations for the SEP analysis in G1 offspring and 75 datasets across 50 iterations in G0 women. In the ACE analysis, results were calculated across 50 imputed datasets (30 iterations) in G1 offspring and 60 imputed datasets (60 iterations) in G0 women.

**Supplementary Note 5: Sensitivity analyses results**

***Complete case***

Full results from the complete case analyses are presented in Supplementary Tables 12-15. In general, the majority of results were consistent with the main results. Few comparisons were directionally the same but statistically weaker than the main analyses, which is reasonable considering the smaller sample size and random variability and does not alter conclusions from the main analyses.

The results from the remaining sensitivity analyses can be found in Supplementary Tables 16-19 and are primarily compared with the complete case results as the most similar analysis in terms of sample size and absence of IPW.

***Parent-specific exposures (SEP only)***

In both the G1 offspring and G0 women, analyses exploring parent-specific SEP exposures were mostly consistent with the main findings or the complete case findings with similar results in both parents. The only notable differences were for the associations between social class and abnormal cycle length and higher education and PMS in the offspring where the associations were only evidenced in the maternal analyses, although the paternal effect estimates were in the same direction. In the G0 women, a slightly larger effect estimate was reported for maternal compared to paternal education regarding the association with cycle length.

***Excluding contraceptive use***

Excluding participants using hormonal contraceptive (OCP in G1 and OCP, IUD, or coil in G0) resulted in few changes to the results beyond some attenuation, which is to be expected given the reduced sample size and does not change any conclusions, particularly as the effect estimates remained in the same direction with similar magnitudes. There were a small number of associations identified in this analyses that were not identified in the main or complete case analyses in the G1 offspring, which may be reflective of chance, collider bias, or true associations previously diluted. When excluding participants using hormonal contraception,

there was evidence for associations between household substance abuse and greater odds of abnormal cycle length, violence between parents and greater odds of PMS-related symptoms, and financial difficulties and lower odds of abnormal cycle length.

***Symptom Severity***

In the G1 offspring participants, we were able to examine severity of pain and heavy or prolonged bleeding based on whether participants sought medical care for that symptom. The effect estimates for SEP exposures tended to be somewhat larger for those who sought medical care compared to those who did not seek medical care for both symptoms. This was also true for the ACE analysis and menstrual pain, where the same associations were evidenced with either similar effect estimates between those who did and did not seek medical care (emotional abuse and violence between parents) or larger effect estimates amongst those who sought medical care (parental mental health problems and parental conviction). The ACE and heavy or prolonged bleeding results were a bit more complex. The same associations were present here as in the complete case analysis, but the effect estimates varied in terms of whether they were smaller (emotional neglect), larger (bullying), or the same (emotional abuse and parental mental health problems) in those who did compared to those who did not seek medical care. Additionally, there was evidence for associations between physical abuse and parental conviction and a greater risk of heavy or prolonged bleeding accompanied by seeking medical care, which was not observed in the main or complete case results.

In G0 women, symptom severity was based on self-report classifications of pain, heavy bleeding, and irregular bleeding. The results for menstrual-related pain were broadly consistent with the main and/or complete case results, with associations between all measures of SEP evidenced except for parental social class, with the largest effect estimates for severe pain. Also, as with the main results, parental SEP, own education, and financial difficulties were not associated with heavy bleeding, regardless of severity. There was, however, evidence that participants own manual occupational social class was associated with a greater risk of reporting severe heavy bleeding. Consistent with the complete case analysis, there was little evidence that parental nor own social class was associated with any severity of irregular bleeding. There was also evidence that financial difficulties were associated with a greater risk of severe irregular bleeding. The relationship between own education and irregular cycles was somewhat complex, with participants educated to lower levels having a lower risk of mild or moderate irregularity, but a higher risk of severe irregularity. In contrast to the complete case findings, there was evidence for an association between lower parental education and a greater risk of severe irregularity. In terms of ACEs, results were similar for menstrual pain, with all associations evidenced in the complete case being observed here with higher effects for severe pain. There was also evidence for associations between sexual abuse and household substance abuse with severe pain that were not observed when examining menstrual pain broadly. No ACEs were associated with heavy bleeding in the complete case analysis, whereas three ACEs were associated with severe heavy bleeding here (physical abuse, emotional neglect, and violence between parents). Finally, sexual abuse (the only ACE associated with irregular bleeding in the complete case analysis) was associated with a greater risk of reporting moderately irregular bleeding only.

***Excluding participants within the first three years since menarche (G1 only)***

When excluding participants within the first three years since menarche, there was stronger evidence for an association between financial difficulties and menstrual pain than was observed in the complete case. Beyond this, the results were similar or slightly attenuated, most likely due to reduced sample size.

***Separating heavy from prolonged bleeding (G1 only)***

When exploring a four-level variable in an attempt to separate heavy from prolonged bleeding in the G1 offspring, the results supported mostly similar conclusions that SEP was not associated with prolonged bleeding and lower parental education was associated with heavy bleeding. In terms of ACEs, the results were consistent regarding no associations between any ACE and prolonged bleeding only. Similarly, all four ACEs that were associated with heavy or prolonged bleeding in the complete case analysis were associated with a higher risk of reporting heavy bleeding only, and three of the four (except emotional abuse) were associated with a higher risk of reporting both heavy and prolonged bleeding. Of the three ACEs associated with both heavy bleeding only and heavy and prolonged bleeding, the effect estimates for the association with emotional neglect and bullying were larger for reporting both symptoms but were similar for the parental mental health associations.

**SUPPLEMENTARY FIGURES**

**Supplementary Figure 1.** Directed acyclic graph (DAG) depicting assumptions about relationships between socioeconomic position (SEP), outcomes, and covariates in G1 offspring (A) and G0 women (A and B).

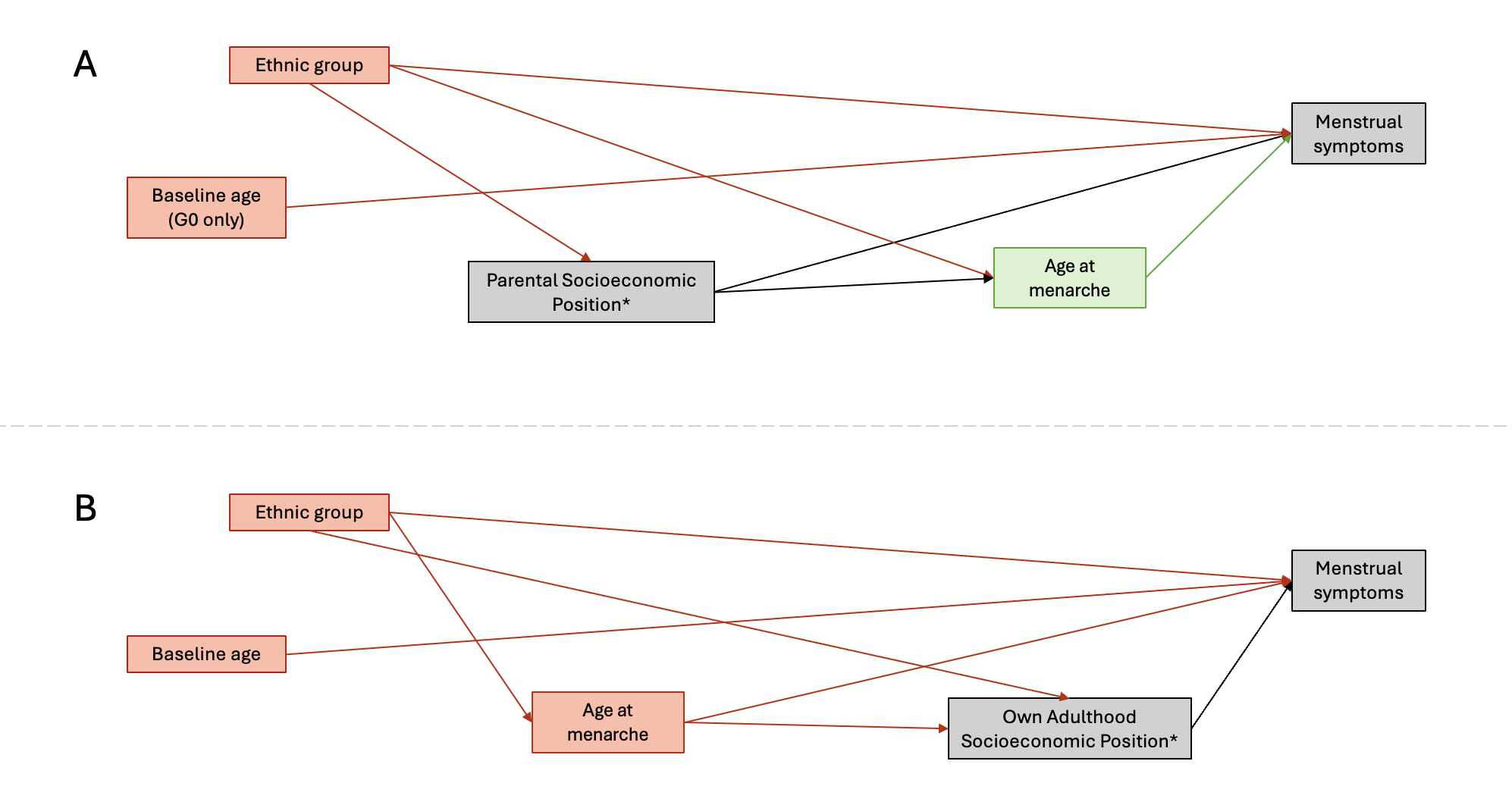

DAG A represents the assumptions in both G1 offspring and G0 women with parental socioeconomic position as the exposure and age at menarche as a mediator. DAG B represents the assumptions in G0 women with own adulthood socioeconomic position as the exposure and age at menarche as a confounder of the association with menstrual symptoms (age at menarche could impact adult SEP or be spuriously associated due to confounding from childhood SEP, so a model with and without age at menarche adjustment has been conducted). Due to the relationship between age and parity, baseline age was adjusted for in G0 women to reduce the age-related variation in menstrual symptom outcomes. *Parental socioeconomic position is defined using highest parental education and occupational social class in both cohorts, as well as financial difficulties during pregnancy for G1 offspring and adulthood socioeconomic position in G0 women is defined using own education, occupational social class, and financial difficulties during pregnancy. Grey boxes represent the association under investigation. Red boxes represent confounders that will be adjusted for. Green boxes (age at menarche) represent mediators of the relationship of interest which are not accounted for.

**Supplementary Figure 2.** Directed acyclic graph (DAG) depicting assumptions about relationships between adverse childhood experiences (ACEs), outcomes, and covariates in G1 offspring (A) and G0 women (A and B).

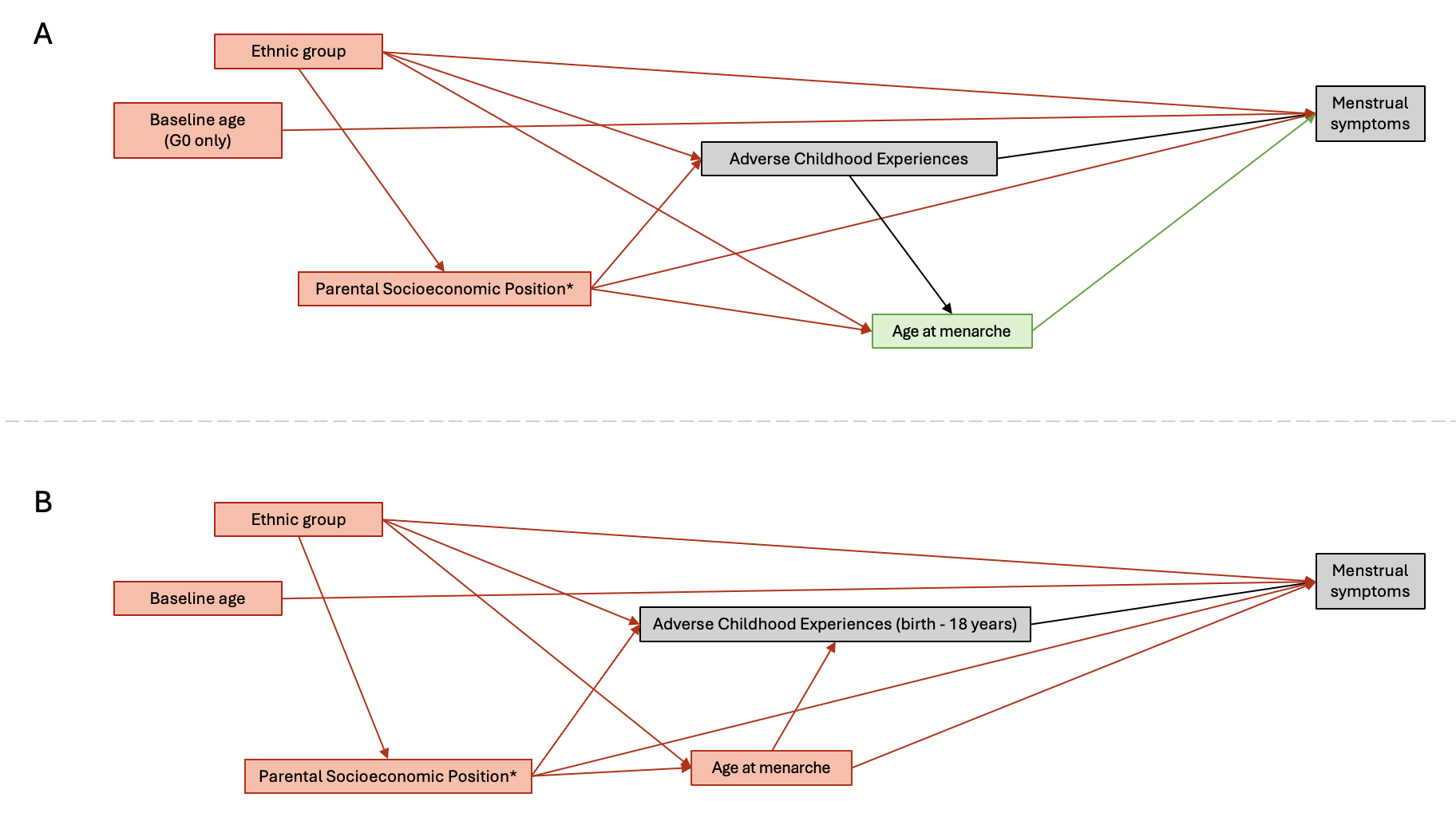

DAG A represents the assumptions in both G1 offspring and G0 women with age at menarche as a mediator of the relationship between ACEs (birth-10 years in G1 offspring and birth-18 years in G0 women) and menstrual symptoms. DAG B represents the alternative assumption in G0 women with age at menarche as a confounder. The relative timing of ACE exposure and age at menarche is unclear in G0 women and therefore a model with and without adjustment for age at menarche has been utilised. Due to the relationship between age and parity, baseline age was adjusted for in G0 women to reduce the age-related variation in menstrual symptom outcomes. *Parental socioeconomic position is defined using highest parental education and occupational social class in both cohorts, as well as financial difficulties during pregnancy for G1 offspring. Grey boxes represent the association under investigation. Red boxes represent confounders that will be adjusted for. Green boxes (age at menarche) represent mediators of the relationship of interest which are not accounted for.

**Supplementary Figure 3.** Flowchart demonstrating how combining MI and IPW allowed a larger, more representative analysis to be conducted using the G1 daughters SEP analysis as an example.

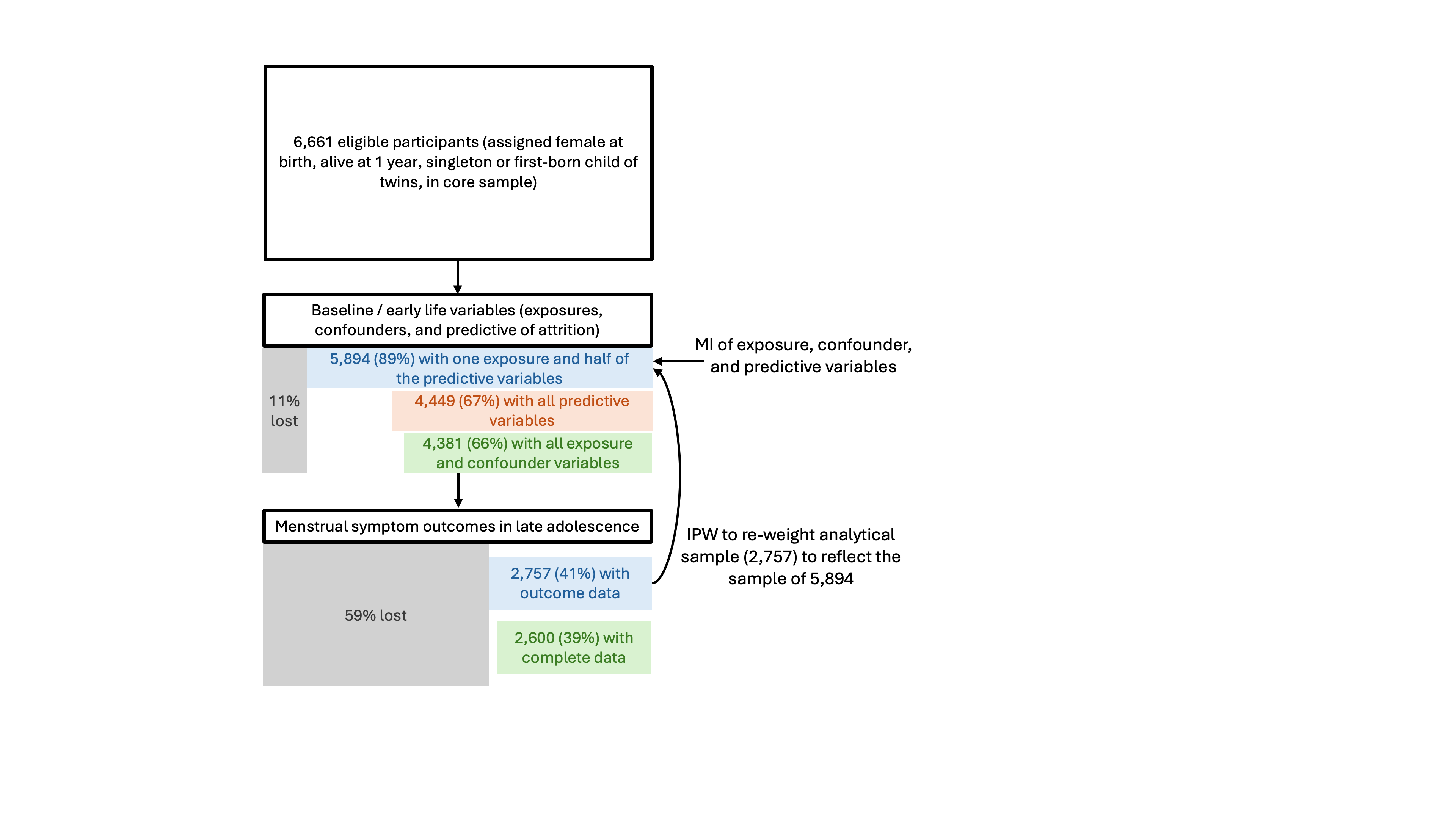

MI of exposure, confounder, and predictive variables was conducted in 89% of the original sample (11% without the minimum amount of non-missing data requirements could not be included). Of these, 2,757 participants (41% of the eligible sample) had non-missing outcome data, constituting the analytical sample, which was re-weighted with IPW to reflect the 89% (5,894 participants). MI in isolation would have enabled analysis in the same sample (41%) but with no weighting; similarly, IPW in isolation would have enabled analysis in 39% of the eligible sample (complete data) and re-weighting to reflect only 6 % (complete data on predictive variables).
